# Implications of Telemedicine on Educational Outcomes and Healthcare Accessibility, Case Study of a Local College in Upstate New York

**DOI:** 10.1101/2020.05.07.20094268

**Authors:** Johnathan Kenyon

**Affiliations:** 118 South Jackson Street Woodbury, NJ 08096, Cazenovia College

## Abstract

**Importance:** There is a dearth of empirical research addressing the healthcare component on the academic performance of college students. It is unknown how discrepancies in healthcare access manifest in college students.

**Objective:** To improve awareness of the current healthcare issues facing college students and find ways to minimize the healthcare gap.

**Design, Setting, and Participants:** Local college students were assessed for health status, usage of the health system, potential factors discouraging the use of health benefits, and the educational outcomes of the students.

**Main Outcomes and Measures:** Educational outcomes associated with differing levels of healthcare access, averages, Pearson’s r, odds ratios and 95% CIs.

**Results:** Nearly 1 in 4 students (26.1% [SE, 3.7%]) reported having inadequate healthcare access. Healthcare accessibility was strongly correlated with academic performance (r = .336 [95% CI, .179-.476]; *P<*.001). Low access students were significantly more likely to report barriers to healthcare (OR = 2.91 [95% CI, .557-5.29]; *P<*.001). Telemedical use corresponded with reduced absences (r = .219 [95% CI, .054-.373]; *P=*.01) and higher ratings on self-health assessments (OR = 1.74 [95% CI, .809-3.75]; *P=*.001). Use of telemedicine did not relate to reports of healthcare barriers (*P*>.99). Though the adoption of telemedicine among college students is staggeringly low, with fewer than 1 in 12 students (7.9% [SE 2.4%]) reporting at least some telemedical usage.

**Conclusions and Relevance:** Over a quarter of college students report inadequate healthcare access despite fewer than 1 in 12 students utilizing telemedicine. Further research is needed to determine the extent to how much the current healthcare norms affect college students, but fostering a pedagogical approach to telemedicine may serve to bridge the healthcare gap.

**Key Points:** *Question:* In what way does healthcare accessibility affect college students?

*Findings:* Many students report inadequate healthcare access, despite having health insurance. The adoption of telemedicine among college students is currently low. Students utilizing telemedicine display improvements across health and academic categories.

*Meaning:* Bolstering the adoption of telemedical health services may help to diminish the healthcare accessibility gap among college students.

## Introduction

Discrepancies in college student healthcare access may exacerbate ignominious educational outcomes. College students could face exorbitant out of pocket costs when seeking healthcare.^1^ Students opting to delay or forgo necessary healthcare could be risking more than their health. Identifying current trends and opening a dialogue for student healthcare could lead to improvements in the quality of life and academic performance of college students.

## Research on Preventive Healthcare

Educational outcomes are inextricably linked to the health of the student. Studies of preventive healthcare in schools K-12 found strong correlations between the availability of preventive health services in schools and academic performance of students K-12.^2^ The critical transition period college represents requires students to become responsible for their own healthcare needs, instead of relying on a third-party to manage their health. This research focuses on investigating the current health climate of local college students and quantifying the relationship between healthcare and academics in order to highlight areas of weakness in healthcare access and propose solutions.

## Methods

### Survey Sample

The target sample was 703 full-time students who were included on Cazenovia College student mailing list (as of April 2020). Survey responses were collected from April 7^th^ through April 12^th^ 2020. The overall response rate was 25.18% (177/703), and the study included only responses with a completion rate greater than 70% (137/177, 77.40%) Informed consent detailing the uses of submitted responses was outlined before students were able to view the survey. Methods for obtaining informed consent and ensuring the privacy of participants were approved and monitored by the institutional review board of Cazenovia College.

### Measures

The survey included four questions relating to the healthcare accessibility status of the student. Two of these questions were used to determine if the student received sufficient healthcare over the past 12 months and the other two questions focused on the healthcare-seeking habits of students, including potential barriers to healthcare access. Factors discouraging students from seeking healthcare are multifactorial. Thus, students may select multiple responses for this question. Two questions]’

>were used to determine the academic performance of the student. One question was used to determine if students used the health insurer from the school or an alternate insurance plan. An additional two questions adapted from a nationwide health survey of college students-^3^ were used to ascertain the general health status of local college students in comparison to college students across the nation.

### Data Analyses

Data collected from the survey results were exported into SPSS. Data were analyzed using SPSS V26. Healthcare usage was analyzed for relationships between self-health assessments, healthcare barriers, and academic performance using binary logistic regression and Pearson’s correlation. Health characteristics of local students were observed for variance with results from a national health survey.

## Results

### Analysis of Healthcare Accessibility

1 in 4 students (26.1% [SE, 3.7%]; Table 1) reported inadequate healthcare access over the last 12 months (for brevity referred to as low access students). Low access students were significantly more likely to report barriers to healthcare (OR = 2.91 [95% CI, .557-5.29]; *P<*.001), with low accessibility showing links to out of pocket expense (OR = 3.85 [95% CI, 1.71-8.69]; *P=*.001), previous unsatisfactory healthcare experiences (OR = 3.58 [95% CI, 1.29-9.93]; *P=*.01), course workload (OR = 2.34 [95% CI, 1.05-5.19]; *P=*.04), and scheduling conflicts (OR = 2.33, [95% CI, 1.05-5.19]; *P=*.04). Students in the bottom quartile reported receiving fewer than one health visit (M = .771 [95% CI, .361-1.180]; *P<*.001) per year. High access was associated with increased self-health assessment scores (r = .267 [95% CI, .104-406]; *P=*.002).

**Table 1.** Complete Survey Results

| **Question** | **Answer Options** | **Students That Answered,**<br>**No./Total No. (%)** |
| --- | --- | --- |
| Do you use the health insurer provided by Cazenovia College? (%) | Yes | 16/136 (11.76) |
|  | No | 120/136 (88.24) |
| In general, how would you rate your overall health? (%)<br>*Adapted from Elflein | Excellent | 19/136 (13.97) |
|  | Very good | 60/136 (44.12) |
|  | Good | 37/136 (27.21) |
|  | Fair | 11/136 (8.09) |
|  | Poor | 8/136 (5.88) |
|  | Don't know | 1/136 (0.74) |
| Within the past 12 months, have you been diagnosed or treated by a health professional for the following? (%)<br>(Select all that apply)<br>*Adapted from Elflein | Sinus infection | 28/137 (20.44) |
|  | Strep throat | 12/137 (8.76) |
|  | Urinary tract infection | 17/137 (12.41) |
|  | Migraine headache | 15/137 (10.95) |
|  | Ear infection | 4/137 (2.92) |
|  | Irritable bowel syndrome | 4/137 (2.92) |
|  | High cholesterol | 1/137 (0.74) |
|  | High blood pressure | 4/137 (2.92) |
|  | Repetitive stress injury (e.g., carpal tunnel syndrome) | 4/137 (2.92) |
|  | Mononucleosis | 3/137 (2.19) |
|  | Diabetes | 2/137 (1.46) |
|  | Endometriosis | 1/137 (0.74) |
|  | HPV | 1/137 (0.74) |
|  | Genital herpes | 1/137 (0.74) |
|  | Gonorrhea | 0/137 (0.00) |
|  | Hepatitis B or C | 0/137 (0.00) |
|  | Pelvic inflammatory disease (PID) | 0/137 (0.00) |
|  | Tuberculosis | 0/137 (0.00) |
|  | HIV | 0/137 (0.00) |
| In the past 12 months, how many classes have you missed because of physical health (e.g., sinus infection, concussion, flu)? (%) | 0 | 33/136 (24.26) |
|  | 1 | 30/136 (22.06) |
|  | 2 | 17/136 (12.50) |
|  | 3 | 13/136 (9.56) |
|  | 4 | 12/136 (8.82) |
|  | 5 | 10/136 (7.35) |
|  | >5 | 21/136 (15.44) |
| What is your approximate GPA? (%) | >3.50 | 67/136 (49.26) |
|  | 3.00 to <3.50 | 37/136 (27.01) |
|  | 2.50 to <3.00 | 20/136 (7.35) |
|  | <2.50 | 1/136 (0.74) |
|  | Prefer not to answer | 11/136 (8.09) |
| In the past 12 months, how many times did you visit a primary care medical professional? (%) | 0 | 9/129 (6.98) |
|  | 1 | 24/129 (18.60) |
|  | 2 | 28/129 (21.71) |
|  | 3 | 25/129 (19.38) |
|  | 4 | 11/129 (8.63) |
|  | 5 | 5/129 (3.88) |
|  | 6 | 7/129 (5.43) |
|  | 7 | 2/129 (1.55) |
|  | >7 | 18/129 (13.95) |
| In the past 12 months, were you able to visit with your primary care medical professional as often as you needed? (%) | Yes | 99/134 (73.88) |
|  | No | 35/134 (26.12) |
| How much time does your primary care medical professional typically spend with you during your visits? (%) | 1 to 5 minutes | 4/133 (3.01) |
|  | 5 to 10 minutes | 32/133 (24.06) |
|  | 10 to 15 minutes | 34/133 (25.56) |
|  | 15 to 20 minutes | 34/133 (25.56) |
|  | >20 minutes | 29/133 (21.80) |
| What, if any, factors are discouraging you from seeking healthcare? (%)<br>(Select all that apply) | Out-of-pocket cost | 44/137 (32.12) |
|  | Scheduling conflicts | 71/137 (51.82) |
|  | Course workload | 46/137 (33.58) |
|  | Out-of-network coverage | 9/137 (6.57) |
|  | Previous unsatisfactory healthcare experience | 18/137 (13.14) |
|  | Severity of health concern | 23/137 (16.79) |
|  | Self-diagnosing | 22/137 (16.06) |
|  | Other | 22/137 (16.06) |
| Do you utilize telemedicine to complete some or all your health visits? (%) | Yes | 10/126 (7.94) |
|  | No | 116/126 (92.06) |

### Analysis of Academic Performance

Self-health assessments show a nearly linear correlation (r = -.502 [95% CI, -.619--.363]; *P<*.001) to the number of classes missed because of illness. Healthcare accessibility was strongly correlated with academic performance (r = .336 [95% CI, .179-.476]; *P<*.001) with low access students showing diminishes in GPA (M = 2.8 [95% CI, 2.5-3.1]; *P<*.001), compared to high access students (M = 3.5 [95% CI, 3.3-3.7]; *P<*.001). Deficiencies in healthcare access corresponded with increases in classes missed due to illness (r = -.348 [95% CI, -.487—.192]; *P<*.001), with high access students reporting fewer absences (M = 2.45 [95% CI, 2.0-2.9]; *P=*.001) than low access students (M = 4.1 [95% CI, 3.1-5.2]; *P=*.001).

### Analysis of Telemedical Usage

Only 1 in 12 students (7.9% [SE, 2.4%]; Table 1) utilize telemedicine for some or all their health needs. Telemedical usage corresponded with shorter interactions with physicians (r = .388 [95% CI, .236-.521]; *P<*.001) with visits typically lasting 4.5 minutes ([95% CI, 1.9-7.1]; *P<*.001). Telemedical savvy students reported fewer illness-related absences (M = .90 [95% CI, .27-1.53]; *P=*.01) than even high access students (r = .219 [95% CI, .054-.373]; *P=*.01). Telemedical users were more likely to report higher ratings on self-health assessment (OR = 1.74 [95% CI, .809-3.75]; *P=*.001) than other students. Usage of telemedicine was unrelated to reports of barriers to healthcare access (*P*>.99) nor was there a relation to telemedical use in either of the health insurance options used by students (*P=*.24).

### Sources of Health Insurance

Approximately 1 in 8 students (11.8% [SE, 2.8%]; Table 1) receive health insurance through the institution. Students sourcing their health insurance from the institution displayed a significantly higher distribution (Mdn = 6 [SD, 2.8]; *P<*.001) of annual health visits than students reporting alternative insurance (Mdn = 2 [SD, 2.3]; *P<*.001) (r = .309 [95% CI, . 150-.453]; *P<*.001). Students receiving insurance through the institution reported greater allotment of time (M = 15 [SD, 6.1]; *P=*.046) during health visits than students reporting alternate insurance (M = 12 [SD, 5.6]; *P=*.046) (r = .171 [95% CI, .004-.329]; *P=*.046). However, these categorizations of health insurance did not relate to healthcare accessibility (*P=*.581).

### Health Characteristics of Local and National College Students

The overall incidence of illness from local students did not significantly differ from students across the nation (Figure 1).^4^ The highest variance between groupings was observed in the sinus infection category. Local college students reported 4.04% (SE 7.52%) more cases of sinus infections than students across the nation (Figure 1). However, the average variance between the two groups across all the categories were 1.2% (Figure 1) remaining within the margin of error.

**Figure 1.**
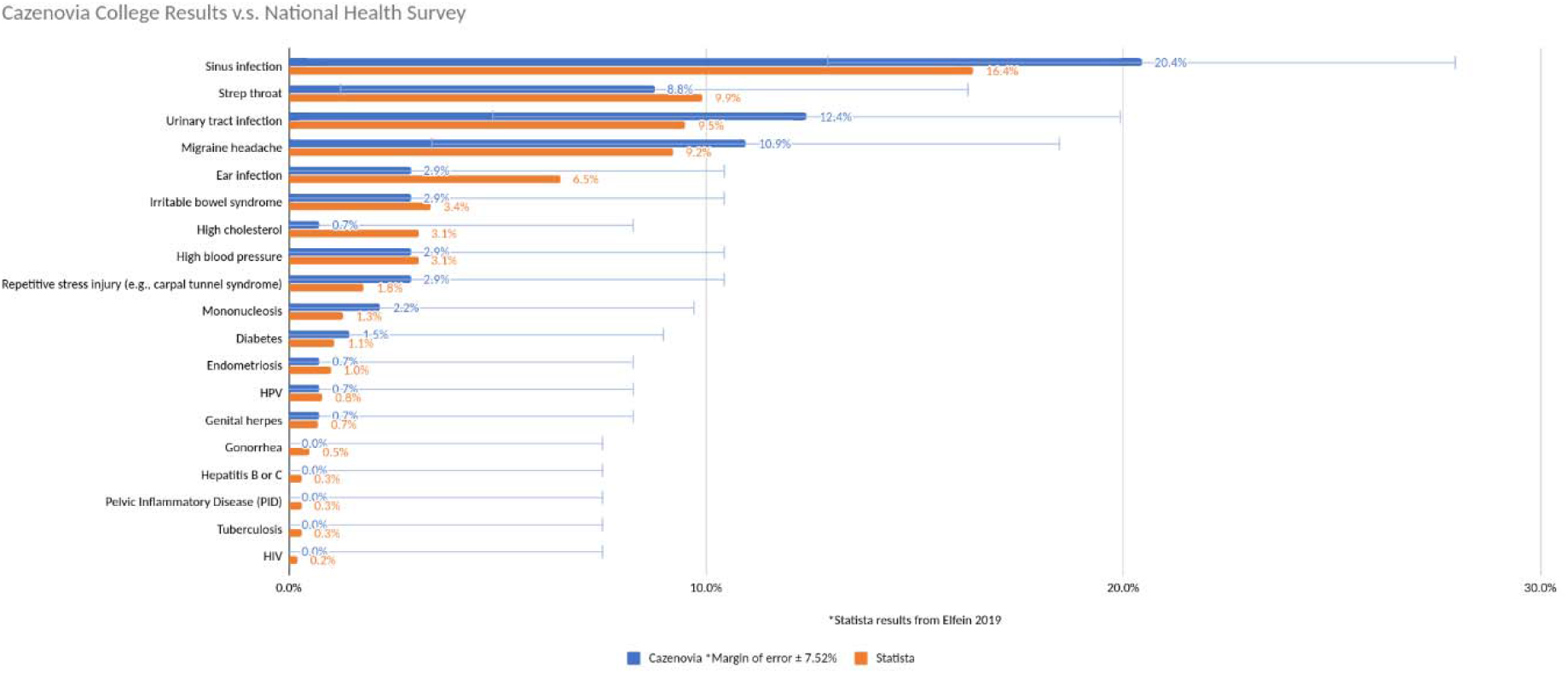
Cazenovia College Health vs Nation-Wide College Student Health Survey

## Discussion

### Health Benefits Utilization

Out of pocket expense discourages healthcare seeking behaviors. This is strongly associated with low healthcare accessibility. Economically vulnerable students would likely benefit the most from improvements in healthcare accessibility. Students that waived the institutional health insurance displayed dramatically fewer health encounters. Could this be motivated by fear of increases to tuition? Healthcare economics are serious considerations for financially vulnerable students, despite healthcare access serving as a strong predictor of educational outcomes.

### Cazenovia College Results Compared with Nation-Wide College Results

The overall health of the local college student population mirrors the results reported from the nation-wide college student health survey. However, the paucity of information regarding the healthcare accessibility of students from other institutions hampers any meaningful comparison. Notably, there is a consistency between local and national health survey results. This raises questions into how healthcare discrepancies manifest in students across the nation, as the location of this study site is in the state with the second lowest rate of uninsured adults.^5^

### Suggestions

Prevalence of telemedicine use among college students is staggeringly low. There simply is an absence of studies detailing the prevalence of telemedical usage among any groups, including college students. Expanding telemedical services in the college student population could conceivably serve as a cost-effective means to improve healthcare accessibility for low access students. Institutions could feasibly offer telemedicine as a free service for students that demonstrate financial/medical need (e.g., low access students).

### Limitations

The shortage of literature regarding the effects of healthcare access on academic performance makes comparison of the results very difficult. Individual health insurance policies are not available for comparison to the institutional health plan. Unfortunately, students in the bottom quartile reported dismal healthcare access. This raises questions regarding their status as insured. For example, students may attempt to evade additional costs to attendance by claiming to be insured. Moreover, the legal mandates on insurance status at the study site make this impossible to tease out. Method of data collection required the active online presence of students during the study timeframe, which occurred shortly after an unexpected transition to online learning. The response rate was likely bolstered by the relevance of the study to the cause of distance learning and the increased presence of online usage of students.

## Conclusion

Over 1 in 4 college students’ healthcare needs are not being met. Telemedical adoption among students remains low, with fewer than 1 in 12 students using telemedicine for any of their healthcare needs. Improving the adoption of telemedicine among college students may help alleviate discrepancies in healthcare accessibility. Therefore, future studies into the healthcare component on educational outcomes should address prevalence of telemedical usage.

## Data Availability

Raw data in excel file is available.

## Acknowledgments

I would like to thank Dr. Livermore for your superb teachings of statistics and how to use statistical tools such as SPSS. The mathematical rigor and education you provided has proven to be invaluable in the writing of this paper.

Also, my sister Marie for providing the technology used to perform the calculations on the data. If not for your selfless act, I would have been unable to complete this assignment.

Finally, I wish to thank Dr. Essary for pushing me to take on the task of conducting my own primary research project. Without your encouragement, I would not have attempted a project this bold - especially as a senior thesis – the capstone of my entire undergraduate academic career. None of this would have been possible without you.

## Notes

### Competing Interest Statement

The authors have declared no competing interest.

### Funding Statement

I received no funding of any type for my research

